# Variation in ambulance pre-alert process and practice: Cross-sectional survey of ambulance clinicians

**DOI:** 10.1101/2023.12.14.23299969

**Authors:** Joanne Coster, Fiona Sampson, Rachel O’Hara, Jaqui Long, Fiona Bell, Steve Goodacre

## Abstract

**Background:** Ambulance clinicians use pre-alerts calls to alert emergency departments (EDs) about the arrival of critically ill patients. We explored ambulance clinician’s views and experiences of pre-alert practice and processes using a national online survey.

**Methods:** Ambulance clinicians involved in pre-alert decision-making were recruited via ambulance trusts and social media to complete an anonymous online survey during May-July 2023. Quantitative data was analysed descriptively using SPSS and text data was analysed thematically to illustrate quantitative findings.

**Results:** We included 1298 valid responses from across 10 ambulance services. Analysis identified variation in practice at all stages of the pre-alert process, including reported frequency of pre-alert (7.1% several times a shift, 14.9% once/twice a month).

Most respondents reported that pre-alerts were delivered directly to the ED but 32.8% reported pre-alerting via an ambulance control room. Personal mobile phones were used to make a pre-alert by 46.8% of respondents, with 30% using ambulance radio. A third of respondents always used mnemonics (e.g. ATMIST/SBAR) but 10.2% reported not using any fixed format.

Guidance used to identify patients for pre-alert varied between clinicians and ambulance service, with local ambulance service guidance most commonly used and 20% stating they never use national guidelines. Respondents reported variable understanding of appropriate conditions for pre-alert and particularly students wanted further guidance on silver trauma and medical pre-alerts.

Only 29% or respondents reported receiving specific pre-alert training and 50% reported never receiving feedback. Fewer than 9% reported always being listened to and having the call taken seriously.

**Conclusion:** We identified variation in pre-alert processes and practice that may result in inconsistent pre-alert practice and challenges for clinicians providing time critical care. Guidance and training on the use of pre-alerts may promote more consistent processes and practices.

**WHAT IS ALREADY KNOWN ON THIS TOPIC:**

⍰ Pre-alerts can enable EDs to prepare for the arrival of a critically ill patient.
⍰ There is variation in local ambulance trust pre-alert guidance, in terms of variation in the conditions suitable for pre-alert and alignment with the ACCE/RCEM pre-alert criteria.

**WHAT THIS STUDY ADDS:**

⍰ The study identifies variation in reported practice in how pre-alerts are delivered across ambulance services and between individual clinicians.
⍰ The study identifies a lack of formal training and feedback around pre-alerts and that a majority of ambulance clinicians would find additional training and feedback useful.

**HOW THIS STUDY MIGHT AFFECT RESEARCH, PRACTICE OR POLICY:**

⍰ Training and guidance in the use of pre-alerts could promote more consistent processes and practices
⍰ Further research is needed to better understand how to improve pre-alert practice and increase consistency.

## Introduction

Pre-alerts are a key part of the emergency care process for critically unwell or injured patients who may require time critical treatments and swift senior clinical review. A pre-alert involves an ambulance clinician or service contacting the ED by telephone ahead of the patient’s arrival to provide information about the patient. This enables the ED to prepare staff and make appropriate space in the department (e.g. resus) for the patient’s arrival. In England over 1 in 10 ambulance conveyances are pre-alerts, however analysis of routine data shows variation in which patients get pre-alerted and in which conditions.^(1)^

Due to inconsistency in pre-alert processes, the Healthcare Safety Investigation Bureau recommended that the Ambulance Association of Chief Executives work with appropriate partners to define and operationalise best practice standards and processes and led to the development of the AACE/RCEM pre-alert guidance in 2020^(2)^ However, there is still significant variation and lack of consistency in pre-alert guidance across different ambulance services, with ambulance service guidance varying from the national AACE/RECEM guidance.^(3)^

Pre-alerts have been shown to result in earlier initiation of time-critical treatments, improved processes and clinical outcomes for patients^(4, 5)^ particularly for conditions such as stroke. There are benefits to care processes and pathways when time critical patients are pre-alerted. For example, timely activation of specific clinical pathways or interventions, safer management of ambulance queues and more informed and effective triage. However, over-use of pre-alerts or perceived inappropriate pre-alerts may result in pre-alerts receiving a reduced response (pre-alert fatigue). Clarity of pre-alert communication is important as communication failures during handover of patient information are recognised as ‘high risk scenario for patient safety’ and can be caused by factors such as not using structured handover processes, absence of formal training and lack of shared understanding.^(6)^ There is some evidence that use of structured communication methods, such as SBAR have been found to improve patient safety.^(7)^

This research forms part of a mixed-method multi-site study to understand how pre-alert decisions are made and communicated and the impact of pre-alerts on receiving EDs and patients. We undertook a survey of ambulance clinicians to identify and explore ambulance clinician views on pre-alert processes and experiences, including decision making and communication. We sought to explore whether there are differences in pre-alert processes at service level and also by different types of ambulance clinicians.

## Methods

We undertook a cross sectional online survey nested with a larger mixed methods study.^(8)^ The survey aimed to explore ambulance clinician’s understanding and experiences of the pre-alert process.

### Patient and public involvement

A PPI group consisting of patients and carers with lived experience of the pre-alert process and representing underserved communities reviewed and commented on the survey questions and process. The study’s PPI co-applicant attended project management meetings where the survey was refined and developed. The PPI group attended a workshop where results were presented and their views and insights on the findings were discussed.

### Sampling and recruitment

We surveyed all Ambulance trusts in England. We recruited ambulance clinicians involved in the pre-alert process via local ambulance trusts. Ambulance trusts used their usual staff research recruitment methods including emails, newsletters, staff facebook groups, posters and advertising on twitter. These differed by site. All participants were required to confirm that they are an ambulance clinician involved in pre-alert decision making prior to completing the survey.

### Mode of administration

The survey was administered online using Qualtrics and was accessed via an online link or QR code, open between 1^st^ May 2023 and 14^th^ July 2023, for a minimum of 6 weeks at each site. Participants were required to confirm their understanding of the study and their consent prior to accessing the full survey. Information including QR code and survey web link to aid participant recruitment was sent to each site once research governance approval had been obtained. The survey was developed to be accessible from a number of different electronic devices, including mobile phones, laptops and tablets. At the end of the survey, participants were given the opportunity to anonymously enter a prize draw to win a £50 voucher with one voucher available per ambulance service.

### The content of the questionnaire

The questionnaire was developed based on issues identified in the literature and preliminary analysis of pre-alert focused interviews with 36 ambulance clinicians across 3 ambulance services. The survey questions explored the pre-alert process from decision to pre-alert to ED response and the survey topic areas are described in Box 1. We collected information on respondent characteristics to explore if there were differences in survey responses at service level and by role in service.

**Box 1:** Survey topics.

**Box 1: Survey topics**
|  |  |
| --- | --- |
| **Section 1: making a pre-alert decision** | ▪ Reasons for making a pre-alert<br>▪ Frequency of making a pre-alert<br>▪ Actions when unsure whether to make a pre-alert<br>▪ Guidance used to aid pre-alert decisions<br>▪ Factors affecting decisions to pre-alert<br>▪ Areas where more pre-alert guidance would be useful |
| **Section 2: Undertaking the pre-alert call** | ▪ Who contacts the receiving ED<br>▪ What device is used to contact the receiving ED<br>▪ Is the pre-alert recorded in the patient notes and if so how is it recorded<br>▪ Learning to make a pre-alert<br>▪ Feedback on pre-alert decisions |
| **Section 3: Communicating with the Emergency Department** | ▪ ED staff responses to pre-alert calls (taking pre-alerts seriously, making appropriate plans in the department, listening without interrupting)<br>▪ Pre-alert format used |
| **Other** | ▪ Anything else to add about pre-alerts |

Survey questions used similar formats throughout and included rating scales, multiple and single choice tick boxes and text boxes to provide additional information.

### Survey pilot

An initial draft of the survey was developed by the research team and piloted with ambulance clinicians from different ambulance services. Analysis of interview data and service level pre-alert policy had identified variation in pre-alert practice and policy, therefore we used the survey pilot to develop a questionnaire that was relevant and inclusive to all ambulance services. There were 13 responses to the survey pilot, which involved ambulance clinicians accessing the survey through the Qualtrics platform and answering the survey questions. Additional feedback on the survey was collated via email. In addition, the survey was also reviewed by each of the ambulance service trusts as part of the local research sign off process and comments about the survey emailed to the study team. This resulted in some changes to the survey, for example, a reduction in the questions included in the survey. The final survey was approved by local ambulance service trusts and the study management group. A copy of the survey is provided as Appendix 1.

The survey did not collect any identifiable information such as name or email address. However, if participants wished to enter the survey prize draw, they selected a link which asked them to enter their email address into a separate form if they wanted to be entered for the prize draw. Information from the prize draw was stored separately and could not be linked to survey responses.

### Analysis

Survey data was collated in the Qualtrics platform and downloaded to SPPS 28 for analysis.^(9)^ We received 266 partial responses (completed <70% of the survey) and these were excluded. Figure 1 describes the survey responses and exclusion process. Variables were cleaned and modified to facilitate analysis. Categorical data were assigned a numerical value and labelled and responses reported at the number and proportion in each category. Continuous data from rating scale answers were reported using the mean, standard deviation and proportion of responses at the scale endpoints. Any missing values were coded as missing.

**Figure 1:**
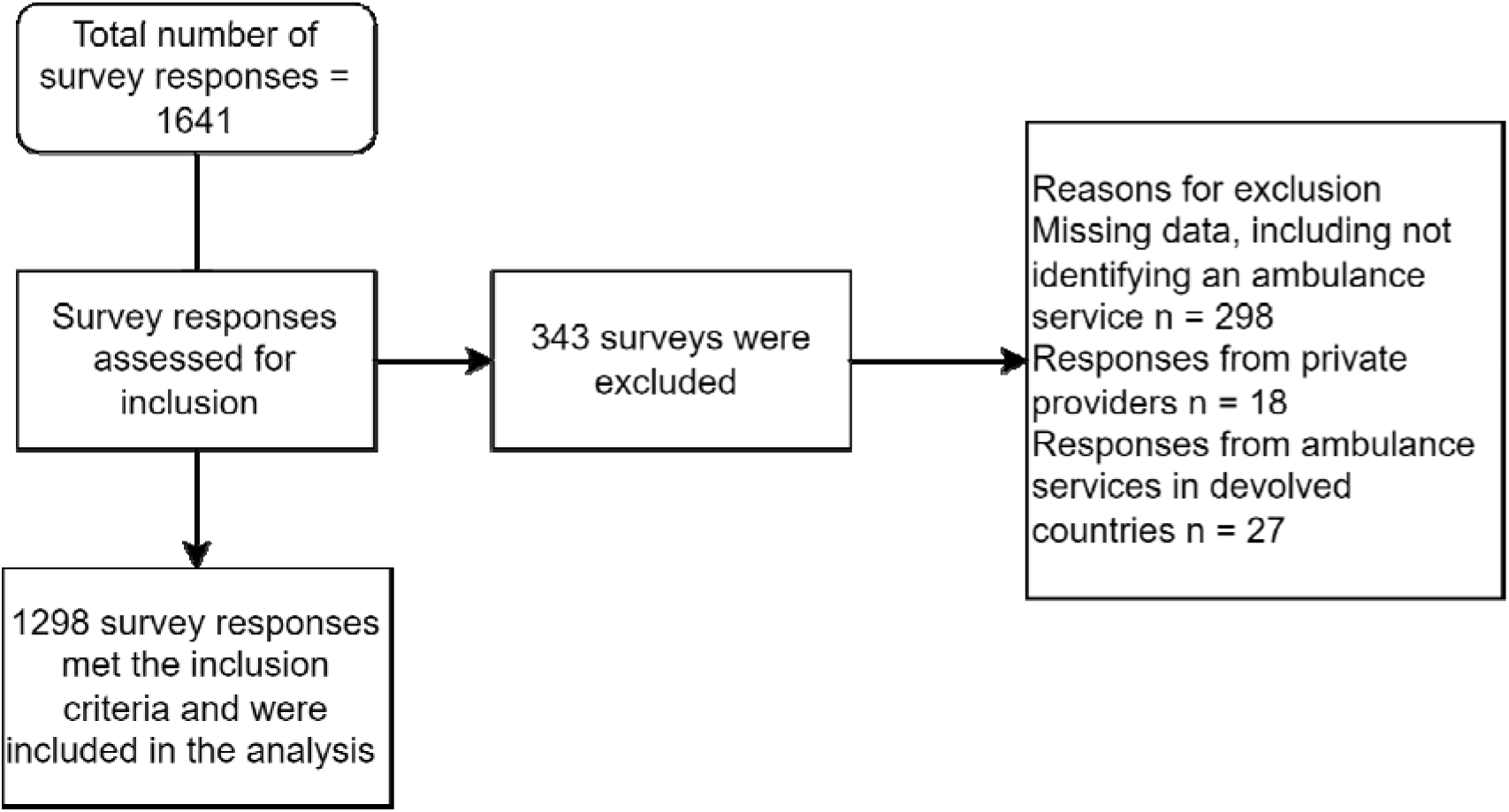
Survey responses and exclusions

A descriptive analysis of the data was undertaken to address the primary aim of describing how ambulance clinicians make pre-alert decisions and undertake pre-alert calls. Variation was explored through subgroup analyses using ambulance service and role in ambulance service variables.

Free text responses were extracted into MicroSoft Excel and coded using a thematic framework developed for the study interviews. Text data was used to further understand the experiences and views of the pre-alert process.

## Results

1641 responses to the survey were received and 343 were excluded (see Figure 1) resulting in 1298 complete responses from 10 English ambulance services included in the analysis. The majority of respondents accessed the survey using the web link, with only 240 respondents using the QR code.

Table 1 describes respondent characteristics. Response rates varied by service. Two services (services 4 and 6) did not have capacity to promote the survey internally resulting in a lower response. Over 50% of the sample were paramedics, with a further 14% being specialist or senior paramedics. Almost half (45%) of the sample had been in their role for >6 years. Most respondents were white males 56.5% (n=734), or white females 36.5% (n=473). Whilst the sample was not diverse in terms of ethnicity, it is broadly representative of people who work as ambulance clinicians in England.^(10)^ This is comparable with NHS workforce data, which reports the proportion of clinically trained ambulance staff as 95.8% white. Comparatively within this survey 94.3% of respondents are white.

**Table 1:**
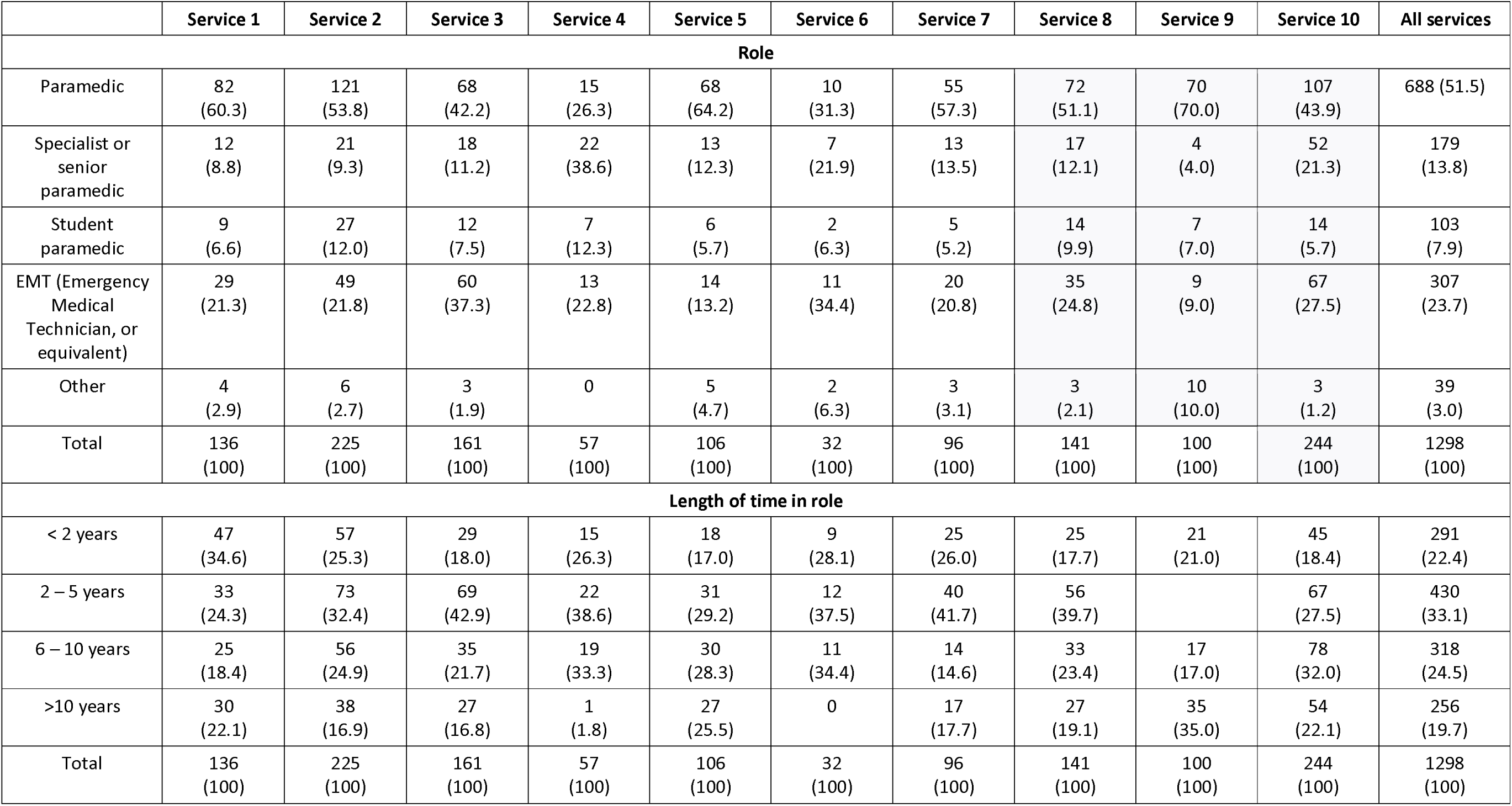
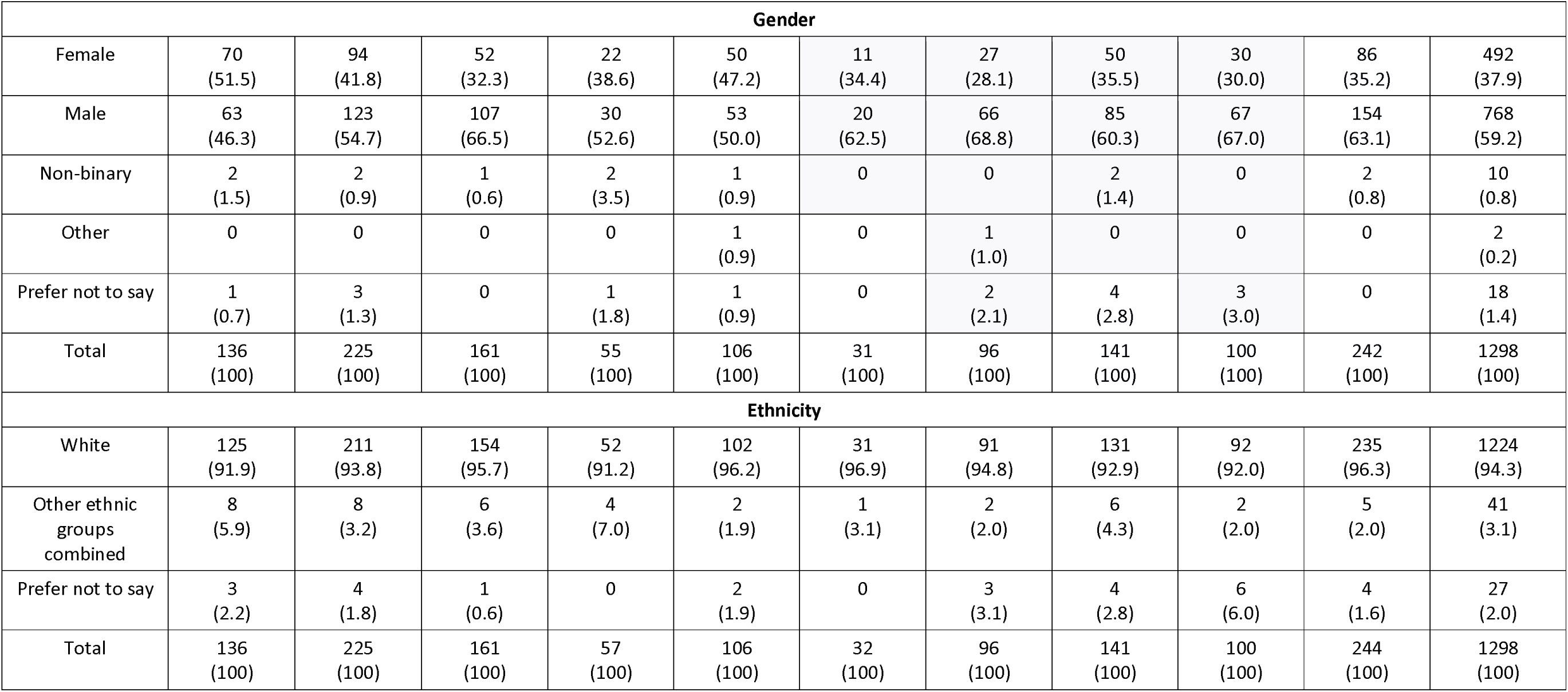
Respondent and workforce characteristics.

### Pre-alert decision making and frequency

Over 80% of respondents reported making a pre-alert either frequently, often or once or twice a week (1061/1298) (Table 4). However, text responses showed that this was difficult to quantify due to the variability of patients seen.

> “Very much pot luck. You can have a run of shifts where every other job is a pre alert and others where you do none.” (Service 2)

Table 2 reports ambulance clinician’s reasons for making pre-alert calls. Clinicians most commonly reported pre-alert calls to be to inform the ED of a deteriorating patient or to make space in resus. There was variation in how often the pre-alert phone was used for advice calls between different services. Text comments identified other reasons for making a pre-alert including; to advise the receiving hospital of additional needs e.g. translation services, mental health, infectious patient; to comply with protocols; to request specific specialists; to advise of violent or difficult to manage patients; Ambulance clinicians also perceived that some patients were too sick to queue and required a different response from the ED, but did not always need resus.

> “The patient doesn’t need resus, but cannot be at the back of an 11 patient queue in the corridor. They need rapid assessment and triage, though not necessarily rapid treatment in resus (Service 1)

**Table 2:** reasons for making a pre-alert call and factors impacting on pre-alert decisions.

|  | Mean<br>(1=Never;<br>5=Always) | SD | Always<br>N (%) | Never<br>N (%) | Paramedics | Specialist/<br>senior<br>paramedics | Student<br>paramedics | EMTs |
| --- | --- | --- | --- | --- | --- | --- | --- | --- |
| **Reason for making a pre-alert** |  |  |  |  |  |  |  |  |
| To inform ED staff of a potentially deteriorating patient | 4.06 | 0.933 | 491<br>(38.2) | 10<br>(0.8) | 4.17 | 3.79 | 4.22 | 3.90 |
| To give the ED time to make space in resus | 3.79 | 1.091 | 401 (31.7) | 31<br>(2.5) | 3.90 | 3.47 | 3.77 | 3.68 |
| To ensure the patient is seen quicker on arrival | 3.73 | 1.177 | 380<br>(31.9) | 73<br>(6.1) | 3.82 | 3.53 | 3.78 | 3.66 |
| For advice about where to take the patient | 2.50 | 1.218 | 55<br>(5.4) | 268<br>(26.5) | 2.16 | 3.15 | 2.45 | 2.80 |
| **Sources of guidance used to help make pre-alert decisions** |  |  |  |  |  |  |  |  |
| JRCALC | 3.49 | 1.13 | 255<br>(19.6) | 59<br>(4.5) | 3.54 | 3.24 | 3.53 | 3.49 |
| Local ambulance trust | 3.75 | 1.06 | 338<br>(26.0) | 36<br>(2.8) | 3.81 | 3.62 | 3.94 | 3.65 |
| Local hospital | 3.10 | 1.25 | 140<br>(10.8) | 149<br>(11.5) | 2.99 | 3.42 | 3.23 | 3.14 |
| ACCE/RCEM | 2.87 | 1.27 | 91<br>(7.0) | 202<br>(20.6) | 2.65 | 3.31 | 2.92 | 3.06 |
| **Factors impacting on pre-alert decision making** |  |  |  |  |  |  |  |  |
| Hospital transporting to | 3.16 | 1.147 | 128<br>(9.9) | 119<br>(9.2) | 3.06 | 3.43 | 3.16 | 3.23 |
| Distance from hospital | 2.94 | 1.21 | 111<br>(8.6) | 162<br>(12.5) | 2.78 | 3.06 | 3.05 | 3.15 |
| Anticipated Queue at the ED | 3.00 | 1.19 | 100<br>(7.7) | 151<br>(11.6) | 2.92 | 3.22 | 2.98 | 3.10 |
| Approaching end of shift | 2.25 | 1.33 | 54<br>(4.2) | 369<br>(28.4) | 1.88 | 2.95 | 2.17 | 2.50 |
| **Physiological criteria or specific conditions that trigger you to make a pre-alert call** |  |  |  |  |  |  |  |  |
| Tachycardia >=131 | 3.54 | 0.99 | 234<br>(18.0) | 19<br>(1.5) | 3.54 | 3.45 | 3.71 | 3.53 |
| Cardiac/Respiratory arrest | 4.54 | 0.90 | 966<br>(74.4) | 2<br>(0.2) | 4.77 | 4.06 | 4.59 | 4.25 |
| Unconscious with a GCS motor score of less than 4 | 4.43 | 0.89 | 829<br>(63.9) | 4<br>(0.3) | 4.62 | 3.98 | 4.49 | 4.20 |
| Respiratory rate =25 | 3.68 | 0.97 | 299<br>(23.0) | 12<br>(0.9) | 3.68 | 3.60 | 3.86 | 3.66 |

The survey explored what ambulance clinicians would do if they were unsure about whether to make a pre-alert call. Table 2 shows that over half stated they would make the pre-alert call anyway with nearly a quarter stating they would call the pre-alert phone to discuss with the ED. Text responses identified a practice of making ‘courtesy calls’ or ‘heads up’ calls.

> “Either make a “courtesy call” to the pre alert phone to give them a heads up you’re coming, this PT could deteriorate but also could be absolutely fine, so when you turn up it’s not a massive surprise if they’re on the edge of deteriorating” (Service 8)

The survey identified that a range of different sources of guidance are used by ambulance clinicians to aid pre-alert decisions. Local ambulance trust guidance was identified as most used and this was consistent across different types of ambulance clinician. There was variation in the use of the national AACE/RCEM guidance, with a fifth of ambulance clinicians never using this guidance, although this varied by service. Text comments identified differences in the guidance and thresholds used by ambulance clinicians for a pre-alert and those used by the ED, which sometimes led to EDs seemingly rejecting a pre-alert or responding dismissively to a pre-alert which met local or national ambulance pre-alert guidance.

> “Our trust uses the RCEM/AACE guidance, but I do feel that sometimes ED scoff/“roll their eyes” even with this. (Service 8)

> “It’s infuriating when following specific guidance which dictates pre alert but finding ED essentially not taking it seriously on your arrival” (Service 5)

> “One of the biggest challenges is ambulance services and hospitals having differing views/policies on what would warrant a pre alert. There needs to be a clear, consistent criteria that both ambulance staff and hospitals follow. Sometimes, I am met by a poor attitude from staff in ED due to them thinking an alert is unnecessary even though it is within my guidance to make the alert.” (Service 2)

### Clinical pathways

We identified variation in pre-alert practice for clinical pathways where there is clear national guidance around making a pre-alert, with three quarters of clinicians reporting always pre-alerting cardiac/respiratory arrest, in comparison with under a quarter alerting for patients with tachycardia of >=131 and respiratory rate of 25.

Variation was identified in relation to pre-alert decision making where there was no condition specific clinical pathway (Table2). Hospital destination had the most impact on pre-alert decision making for all staff groups. Approaching end of shift was considered to have the least impact on pre-alert decision making.

Text comments identified tensions in trying to balance ED variation around what should be pre-alerted.

> “Every ED seems to be different and there is a huge variation even between staff within the same ED to pre-alerted patients, which makes it seem like whatever you do/don’t pre-alert, you are invariably in the wrong (according to them). I also think it is difficult with conditions such as sepsis, where if you follow the trust policy and pre-alert, you will get eye-rolled and no bed/quicker treatment/response for the patient, so it almost feels embarrassing doing the pre-alert but then it feels like there’s the risk of getting into “trouble” from the ambulance trust if you don’t stick to the policy they have written.” (Service 10)

> “The challenge is not just the pre-alert process but also navigating which types of patients which hospitals want pre-alerted or not. For instance, in my area, one hospital has a fractured NOF pathway and want a pre-alert, but the other hospital doesn’t, so don’t want a pre-alert for fractured NOF patients. The local TUs will often tell you that you should take a trauma patient to an MTC during the pre-alert call, despite the patient not meeting the local decision-tree criteria for MTC” (Service 7)

Respondents indicated that in most areas, more guidance would be well received, particularly in silver trauma (65% would like more guidance), and medical pre-alerts generally where 57% of ambulance clinicians would like further guidance (Table 3).

**Table 3:** Types of patients where further guidance would be welcomed, by job role type.

|  | % Yes |  |  |  |  |
| --- | --- | --- | --- | --- | --- |
| Further guidance | Paramedic | Specialist | Student | EMT | All staff |
| Trauma generally | 346<br>(51.8) | 56<br>(31.3) | 66<br>(64.1) | 143<br>(46.6) | 639<br>(49.2) |
| Silver trauma | 456<br>(68.3) | 91<br>(50.8) | 76<br>(73.8) | 187<br>(60.9) | 842<br>(64.9) |
| Medical pre-alerts generally | 388<br>(58.1) | 80<br>(44.7) | 62<br>(60.2) | 188<br>(61.2) | 744<br>(57.3) |
| Sepsis | 273<br>(40.9) | 66<br>(36.9) | 51<br>(49.5) | 130<br>(42.3) | 541<br>(41.7) |
| Respiratory | 271<br>(40.6) | 48<br>(26.8) | 61<br>(59.2) | 122<br>(39.7) | 524<br>(40.4) |

### Pre-alert calls and processes

Pre-alert practice by service is reported in Table 4:

**Table 4:** Pre-alert practice by ambulance service.

|  | Service 1 | Service 2 | Service 3 | Service 4 | Service 5 | Service 6 | Service 7 | Service 8 | Service 9 | Service 10 | All services |
| --- | --- | --- | --- | --- | --- | --- | --- | --- | --- | --- | --- |
| **Pre-alert frequency** |  |  |  |  |  |  |  |  |  |  |  |
| Frequently e.g. several times a shift | 17<br>(12.5) | 17<br>(7.6) | 8<br>(5.0) | 5<br>(8.8) | 5<br>(4.7) | 4<br>(12.5) | 5<br>(5.2) | 2<br>(1.4) | 2<br>(2.0) | 27<br>(11.1) | 92<br>(7.1) |
| Often e.g once a shift | 39<br>(28.7) | 56<br>(24.9) | 48<br>(29.8) | 13<br>(22.8) | 28<br>(26.4) | 8<br>(25.0) | 25<br>(26.0) | 32<br>(22.7) | 13<br>(13.0) | 75<br>(30.7) | 337<br>(26.0) |
| Sometimes e.g. once or twice per week | 58<br>(42.6) | 118<br>(52.4) | 82<br>(50.9) | 24<br>(42.1) | 51<br>(48.1) | 14<br>(43.8) | 50<br>(52.1) | 75<br>(53.2) | 50<br>(50.0) | 110 (45.0) | 632<br>(48.7) |
| Infrequently e.g. once or twice per month | 15<br>(11.0) | 23<br>(10.2) | 20<br>(12.4) | 14<br>(24.6) | 21<br>(19.8) | 6<br>(18.8) | 10<br>(10.4) | 28<br>(19.9) | 31<br>(31.0) | 26<br>(10.7) | 194<br>(14.9) |
| Other | 7<br>(5.1) | 10<br>(4.4) | 3<br>(1.9) | 1<br>(1.8) | 1<br>(0.9) | 0 | 5<br>(5.2) | 3<br>(2.1) | 4<br>(4.0) | 5<br>(2.0) | 39<br>(3.1) |
| **Who contacts the ED** |  |  |  |  |  |  |  |  |  |  |  |
| Crew on scene | 89<br>(65.4) | 108<br>(48.0) | 126<br>(78.3) | 14<br>(24.6) | 10<br>(9.4) | 15<br>(46.9) | 80<br>(83.3) | 122<br>(82.5) | 91<br>(91.0) | 56<br>(23.0) | 711<br>(54.8) |
| Ambulance control centre | 26<br>(19.1) | 29<br>(12.9) | 32<br>(19.9) | 37<br>(64.9) | 94<br>(88.7) | 14<br>(43.8) | 13<br>(13.5) | 16<br>(11.3) | 5<br>(5.0) | 160<br>(65.6) | 426<br>(32.8) |
| Someone else in the ambulance service | 0 | 1<br>(0.4) | 0 | 0 | 1<br>(0.9) | 0 | 0 | 1<br>(0.7) | 1<br>(1.0) | 12<br>(4.9) | 16<br>(1.2) |
| Crew on scene medical alerts and trauma desk for trauma alerts | 20<br>(14.7) | 84<br>(37.3) | 3<br>(1.9) | 6<br>(10.5) | 1<br>(0.9) | 2<br>(6.3) | 1<br>(1.0) | 2<br>(1.4) | 3<br>(3.0) | 16<br>(6.6) | 138<br>(10.6) |
| **Device usually used to make the call** |  |  |  |  |  |  |  |  |  |  |  |
| Personal mobile | 68<br>(50.0) | 138<br>(61.3) | 125<br>(77.6) | 23<br>(40.4) | 19<br>(17.9) | 12<br>(37.5) | 11<br>(11.5) | 110<br>(78.0) | 82<br>(82.8) | 19<br>(7.9) | 607<br>(46.8) |
| Work mobile | 49<br>(36.0) | 19<br>(8.4) | 26<br>(16.1) | 21<br>(36.8) | 13<br>(12.3) | 15<br>(46.9) | 8<br>(8.3) | 19<br>(13.5) | 3<br>(3.0) | 55<br>(22.8) | 228<br>(17.6) |
| Ambulance radio | 15 | 66 | 10 | 11 | 58 | 4 | 72 | 11 | 10 | 133 | 390 |
|  | (11.0) | (29.3) | (6.2) | (19.3) | (54.7) | (12.5) | (75.0) | (7.8) | (10.0) | (55.2) | (30.0) |
| Other | 3<br>(2.2) | 1<br>(0.4) | 0 | 0 | 1<br>(0.9) | 0 | 3<br>(3.1) | 0 | 4<br>(4.0) | 1<br>(0.4) | 62<br>(4.8) |
| **How pre-alert is recorded** |  |  |  |  |  |  |  |  |  |  |  |
| Free text only | 11<br>(8.1) | 27<br>(12.0) | 28<br>(17.4) | 13<br>(22.8%) | 15<br>(14.2) | 4<br>(12.5) | 5<br>(5.2) | 10<br>(7.1) | 18<br>(18.0) | 34<br>(13.9) | 165<br>(12.7) |
| Free text plus tick box for pre-alert | 113<br>(83.1) | 183<br>(81.3) | 110<br>(68.3) | 39<br>(68.4) | 75<br>(70.8) | 24<br>(75.0) | 52<br>(54.2) | 99<br>(70.2) | 70<br>(70.0) | 164<br>(67.2) | 929<br>(71.6) |
| Tick box only | 7<br>(5.1) | 9<br>(4.0) | 19<br>(11.8) | 4<br>(7.0) | 15<br>(14.2) | 2<br>(6.3) | 33<br>(34.4) | 28<br>(19.9) | 9<br>(9.0) | 44<br>(18.0) | 170<br>(13.1) |
| Other | 3<br>(2.2) | 4<br>(1.8) | 4<br>(2.5) | 0 | 0 | 1<br>(3.1) | 4<br>(4.2) | 4<br>(2.8) | 3<br>(3.0) | 2<br>(0.8) | 25<br>(1.9) |
| **Given specific training on how to pre-alert** |  |  |  |  |  |  |  |  |  |  |  |
| Yes | 44<br>(32.4) | 43<br>(19.1) | 36<br>(22.4) | 15<br>(26.3) | 28<br>(26.4) | 5<br>(15.6) | 17<br>(17.7) | 46<br>(32.6) | 31<br>(31.0) | 102<br>(41.8) | 367<br>(28.3) |
| No | 91<br>(66.9) | 181<br>(80.4) | 125<br>(77.6) | 41<br>(71.9) | 78<br>(73.6) | 26<br>(81.3) | 77<br>(80.2) | 95<br>(67.4) | 69<br>(69.0) | 140<br>(57.4) | 854<br>(65.8) |
| **Feedback (from ED or ambulance service)** |  |  |  |  |  |  |  |  |  |  |  |
| Yes | 77<br>(56.6) | 123<br>(54.7) | 88<br>(54.7) | 50<br>(87.7) | 36<br>(34.0) | 22<br>(68.8) | 43<br>(44.8) | 68<br>(48.2) | 44<br>(44.0) | 144<br>(59.0) |  |
| No | 58<br>(42.6) | 101<br>(44.9) | 73<br>(45.3) | 7<br>(12.3) | 69<br>(65.1) | 10<br>(31.3) | 51<br>(53.1) | 73<br>(51.8) | 56<br>(56.0) | 100<br>(41.0) |  |

Variation in practice in the pre-alert call to the ED was mostly service based. In most services the common practice was for the ambulance clinician on scene to make the call to the ED (54.8%; 711/1298) whereas in some services standard practice was for the ambulance clinician on scene to phone through to the ambulance control desk, who would then call the ED pre-alert phone and pass on the information. Practice was sometimes different for medical and trauma calls.

Variation in how pre-alert calls were made was identified and this varied by service (Table 4). Ambulance radios tended to be used infrequently at most services, with most calls made using personal mobile phones. Most respondents reported always recording the pre-alert in the patient notes and using a tick box plus free text, however some variation by service was observed (Table 4).

### Learning how to make a pre-alert

Most survey respondents reported they had not received any specific training on how to make a pre-alert call (65.8%; 854/1298). Other, more informal training methods, were used, such as 59.2% (769/1298) reported learning from a mentor or senior colleague; 58.6% reported learning as they went along/on the job (761/1298); and 20.6% reported learning from written guidelines (267/1298).

Most staff (53.5% 695/1298) reported that they had never received feedback on their pre-alert decisions from either EDs or their ambulance service and this was consistent across most different services. Text comments highlighted the perceived usefulness of feedback, but cautioned that feedback was very often given negatively for a perceived wrong pre-alert decision.

> I was questioned by a clinician receiving the pre-alert on why I was pre-alerting a patient into hospital, despite a genuine clinical concern from ourselves for the patient. The person on the phone stated she thought it was an inappropriate pre-alert. (Service 2)

> As a graduate paramedic I received helpful feedback on every pre-alert I made as a student but I have received no feedback as a qualified paramedic. (Service 8)

> The majority of the time I worry about pre-alerting too much and then worry about making pre-alert calls for pts I am unsure about. A feedback system would be greatly appreciated, as I have never received formal feedback from a hospital. I tend to base my pre-alert decisions on my own clinical judgement (need, observations, intervention, overall clinical picture, ongoing care, formal pathways etc.) in the hopes that this is appropriate. (Service 7)

> How the hospital receives the alert definitely has an impact on what I alert. (Service 8)

### Communication with the ED

9% of ambulance clinicians felt that ED clinicians always listen and take the call seriously, always listen without interrupting and always make appropriate arrangements in the ED. There was little variation by role reported, however student paramedics reported experiencing more interruptions and senior paramedics had the highest ratings for being listened to and taking the call seriously (see table 5).

> Often interrupted or questioned about my decision to pre-alert which takes time away from patient care. (Service 7)

**Table 5:** communication with the ED.

|  | Mean<br>(1=Never;<br>5=Always) | SD | Always<br>N (%) | Never<br>N (%) | Paramedics | Specialist/<br>senior<br>paramedics | Student<br>paramedics | EMTs |
| --- | --- | --- | --- | --- | --- | --- | --- | --- |
| **When making a pre-alert call to the ED, do you feel that ED staff** |  |  |  |  |  |  |  |  |
| Listen to you and take the call seriously | 3.31 | .90 | 114<br>(8.8) | 15<br>(1.2) | 3.24 | 3.53 | 3.24 | 3.36 |
| Listen without interrupting | 3.09 | 1.04 | 115<br>(8.9) | 59<br>(4.5) | 3.04 | 3.11 | 2.83 | 3.26 |
| Make appropriate arrangements in the ED | 3.21 | 0.94 | 111<br>(8.6) | 25<br>(1.9) | 3.16 | 3.38 | 3.20 | 3.20 |
| **When you phone the ED, what format do you follow?** |  |  |  |  |  |  |  |  |
| Use a predefined format e.g. ATMIST, ASHICE, SBAR | 4.00 | 0.98 | 464<br>(35.7) | 16<br>(1.2) | 4.12 | 3.74 | 4.04 | 3.85 |
| Use a *different* pre-defined format, please state | 2.56 | 1.31 | 26<br>(2.0) | 129<br>(9.9) | 2.20 | 3.16 | 2.61 | 2.73 |
| Use the format that the receiving ED uses | 2.92 | 1.31 | 90<br>(6.9) | 170<br>(13.1) | 2.70 | 3.24 | 3.14 | 3.07 |
| Provide observations but don't follow a fixed format | 3.16 | 1.24 | 133<br>(10.2) | 111<br>(8.6) | 3.08 | 3.36 | 3.27 | 2.53 |

> Ed staff often interrupt and do not fully listen and can sound dismissive (Service 10)

> ED staff often lack insight into the fact we have very little bandwidth for the prealert. Often a paramedic is managing an acutely unwell patient and ED staff forget this. ED staff often interrupt and ask questions that can be better answered at handover or simple are not relevant at that time. (Service 2)

> The seniority of the staff member who picks up the phone seems directly linked to how much they interrupt the pre-alert. I.e. a doctor will often just listen, a nurse will interrupt to fit the information in the order they are running through the list their side, which may differ to how the handover is being given. It is easier to pre-alert to a member of hospital staff that you already know because they trust your clinical judgement, as opposed to someone who does not know you. (Service 6)

The format of the communication with the ED varied overall and also by staff type. A third of ambulance clinicians reported always using a fixed format (35.7%; 464/1298), however 1 in 10 reported always providing observations but not following a fixed format (10.2%; 133/1298). Specialist/senior paramedics had slightly lower ratings for using a fixed format and higher ratings for using the format that the receiving ED uses and providing observations but not following a fixed format.

> “We serve several hospital[s] in my area - each hospital appears to have different pre-alert rules - this would determine which hospital receives a pre alert or not” – (Service 1)

> “We have a particular hospital that is notorious for not taking pre-alerts seriously. Last week I pre-alerted a patient with a NEWS 2 of 13 - red flag sepsis & reduced GCS. We had a travel time of 20 minutes yet the p/t was not placed into resus because ‘there are no nurses to watch him’ No Doctor had been informed & the p/t placed onto a normal handover bed in ED where he deteriorated & was moved into resus 20 minutes after we arrived.” (Service 3)

> “This is very frustrating & makes us wonder why we bother making a call. (Service 1) the destination is a key decision maker, some hospitals are better than others when taking pre alerts, some turn into a lengthy unnecessary conversation. As well the level of incivility experienced over the phone and on handover influence a decision to pre alert or not” (Service 1)

## Discussion

### Summary of key findings

Participants reported a lot of variation in their pre-alert practice and the ED response. Some of this variation is service based, most notably, who makes the call to the ED (crew on scene or clinical hub) and service-based variation in guidance or use of checklists. Fewer than one in ten ambulance clinicians perceived that their pre-alert calls were always listened to without interruption, taken seriously, or that appropriate arrangements were made in the ED. Participants reported tensions in trying to balance local ED policy variation around what should be pre-alerted with local ambulance guidance. This challenge may be exacerbated by the range of different types of ambulance pre-alert guidance that were reported as used and reported low usage of the national AACE/RCEM guidance. Whilst national guidance on pre-alerts for Cardiac/Respiratory arrest was always followed by a majority of ambulance clinicians, it was not always followed in a quarter of cases. Survey respondents reported that access to training and feedback are lacking. Most ambulance clinicians reported not receiving any specific training on how to make a pre-alert call and over half stated they had never received feedback about pre-alert decisions.

### Comparison with other literature

Literature exploring how pre-alerts are undertaken and used is limited. Our survey identified differential understanding of which conditions should be pre-alerted and variation in pre-alert rates. Boyd et al identified significant differences in pre-alert guidance available between ambulance services which may explain this variation. Similarly Pilbery et al identified differences in pre-alert rates between ambulance services.^(1)^

Poor communication during handover of information is identified as impacting on patient safety and the timeliness of transfers of care.^(11)^ The survey identified that the use of structure formats varied, and that even where structured formats were used these may not be the same, and that pre-alert communication was often interrupted or not acted upon. Using structured formats, where each person uses the same format to communicate agreed content is thought to improve communication and positively impact on quality of care.^(12)^

Findings related to lack of feedback about pre-alert decisions are mirrored in other countries and health systems. A US survey identified 45.5% of EMS clinicians had not received feedback within a 30 day period^(13)^ and in Canada feedback was identified as not being part of routine practice.^(14)^ A qualitative UK study identified EMS professionals had a ‘strong desire for feedback’ and multiple benefits of constructive feedback including supporting professional development and improving patient care.^(15)^ The beneficial impact of feedback on care processes that form part of the pre-alert processes, such as improving clinical decision making, protocol adherence and documentation was identified in a systematic review.^(16)^

Two thirds of ambulance clinicians were in favour of further guidance about silver trauma. Older trauma patients have complex presentations and benefit from early review from a geriatrician.^(17)^ Therefore additional guidance for prehospital clinicians may facilitate earlier identification and therefore treatment of silver trauma.

### Limitations

Whilst online survey methods have limitations in relation to response rate and potential bias^(18)^, using this method allowed us to gain a national view of pre-alert practice. We obtained a response from 1298 respondents. The high number of responses and engagement shows that this is a salient issue to ambulance clinicians. Response numbers varied between sites, partly due to differences in site recruitment strategies, but is representative of the skill mix, and diversity of ambulance clinicians.

There is potential for response bias, with respondents having stronger views or more negative experiences being more likely to respond. We attempted to counteract non-response bias through engaging with ambulance clinicians to develop the survey questions and formats, developing a survey that was quick and easy to fill in from mobile devices, providing incentives for completion and using a survey pilot to pre-test the questions and ensure their appropriateness to each ambulance service.

Internal validity of survey data can be affected by multiple survey submissions.^(19)^ We recruited via ambulance service research departments, checked IP addresses for duplicates and assessed responses for similarity to counter this. We also ensured that survey completion progress was saved so that returning to complete the questionnaire did not result in the completion of another questionnaire. There was some variation in the number of responses from each site. This was partly due to 2 ambulance services not having the capacity to promote the survey. Despite this, eight trusts each returned over 95 responses.

Whilst the findings reflect what the participants reported rather than what happened, they align with routine data showing that variation in practice that is unexplained by clinical need.^(20)^ Some variation in practice may be due to service level variation. For example, pre-alert recording in the patient notes may be impacted by mandated information recording policies. At least two services had pre-alert systems where pre-alert information was passed to EDs by the ambulance control room, rather than the crew on scene.

### Implications of the results for policy and practice

Wide reported variation in pre-alert policies at ambulance and ED service level shows that there is scope to develop pre-alert policies that are more aligned between services and to embed national policy. Co-producing and embedding national policy within local ambulance and hospital trust and developing enhanced guidance for pre-alerting silver trauma or medical pre-alerts in general may help to increase consistency.

Feedback on pre-alert decision-making was highlighted in the survey as something that ambulance clinicians would generally welcome. However, design and development of feedback mechanisms that do not place additional workload on already busy staff is required.

## Conclusion

We identified wide variation in pre-alert practice and this was partly due to variation in pre-alert policies at ED and ambulance service level. Variation can result in challenges for clinicians involved in pre-alerts at a time when they are caring for time critical patients. Introduction of training and feedback may lead to opportunities for learning and improving pre-alert practice at individual clinician and service levels.

## Data Availability

All data produced in the present work are contained in the manuscript

## Additional information

## Disclaimer

The views expressed are those of the author(s) and not necessarily those of the NIHR or the Department of Health and Social Care.

## Authors’ contributions

JC conceived and designed the study and led this aspect of work, including development of the survey, data collection and analysis. FS conceived, designed. led and oversaw the study, and actively contributed to development of survey materials, the analysis and interpretation of findings. RO and JL contributed to the development and piloting of the questionnaire and the interpretation of findings. JC led the data analysis and drafted the paper. FB conceived and designed the study and contributed to the development of survey materials and recruitment of participants. JL, FS, FB and SG contributed to the data analysis and findings. All authors read drafts of the manuscript and approved the final version. JC is guarantor for the paper.

## Acknowledgements

The authors would like to thank all research participants and ambulance services who helped to recruit participants. We are also grateful for the input of other members of the study team, our advisory group and our patient and public involvement representatives/group. Thanks to Marc Chattle for clerical support.

Thank you to the National Ambulance Research Steering Group (NARSG) for reviewing the research and supporting the research opportunity.

Thank you to the ambulance clinicians and research staff who reviewed the survey and took part in the survey pilot.

## Data Sharing

The data generated for this study is in the form of confidential transcripts of interviews that are not available for sharing. Participants consented for anonymised quotations to be shared but did not consent to share the full transcripts.

## Patient and Public involvement

Patient and public involvement (PPI) representatives contributed to the design and conduct of this study through input at PPI and project management group meetings. Preliminary findings were presented and discussed at a PPI event for the study.

## Ethical and information governance considerations

The survey research plan received ethical approval from NHS ethics North East - Newcastle & North Tyneside 2 Research Ethics Committee 21/NE/0132 and the piloted version of the survey received ethics approval by the same ethics committee as amendment number 7.

